# Digital system augmented by artificial intelligence to interpret bone marrow samples for hematological disease diagnosis

**DOI:** 10.1101/2022.08.30.22279373

**Authors:** David Bermejo-Peláez, Sandra Rueda Charro, María García Roa, Roberto Trelles-Martínez, Alejandro Bobes-Fernández, Marta Hidalgo Soto, Roberto García-Vicente, María Luz Morales, Alba Rodríguez-García, Alejandra Ortiz-Ruiz, Alberto Blanco Sánchez, Adriana Mousa Urbina, Elisa Álamo, Lin Lin, Elena Dacal, Daniel Cuadrado, María Postigo, Alexander Vladimirov, Jaime Garcia-Villena, Andrés Santos, Maria Jesús Ledesma-Carbayo, Rosa Ayala Díaz, Joaquín Martínez-López, María Linares, Miguel Luengo-Oroz

## Abstract

Analysis of bone marrow aspirates (BMA) is an essential step in the diagnosis of hematological disorders. This analysis is usually performed based on visual examination of the samples under a conventional optical microscope, which involves a labor-intensive process, limited by clinical experience and subject to high observer variability. In this work, we present a comprehensive digital system that enables BMA analysis for cell type counting and differentiation in an efficient and objective manner. This system not only provides an accessible and simple method to digitize, store and analyze BMA samples remotely, but is also supported by an artificial intelligence (AI) pipeline that accelerates the differential cell counting (DCC) process and reduces inter-observer variability. It has been designed to integrate AI algorithms with the daily clinical routine and can be used in any regular hospital workflow.

## 1. Introduction

The analysis of Bone Marrow Aspirate (BMA) samples is a common and essential process for several hematological diseases (acute leukemias, plasma cell disorders, myelodysplastic syndromes, etc.)^1,2^. Every day, thousands of hematologists around the world perform BMA analysis, and despite current technological advances, they still heavily rely on traditional optical microscopes and their own clinical expertise.

As has been stated in several studies, BMA analysis shows a high level of interobserver variability (either in zone selection to analyze the sample or when classifying and counting hematopoietic stem cell lineages)^3–6^. Furthermore, the International Council for Standardization in Haematology (ICSH) guideline for the standardization of bone marrow specimens and reports suggests extending the number of cells to be counted to more than 500, or even comparing the results with those of another sample and asking another observer to evaluate the sample independently, especially when a disease is suspected^7^. In short, the differential cell count (DCC) in BMA samples has proved to be a time-consuming and error-prone procedure^8^ that requires a hematologist highly specialized in cytology to ensure reliable results^9^.

So far, a quite limited number of tools and devices have been developed and validated to assist professionals during the DCC process and to increase their efficiency by leveraging artificial intelligence (AI). An extended literature review can be found in supplementary **Table 1S**.

On the one hand, many authors have proposed the use of commercially available slide scanners or digital cameras attached to optical microscopes to obtain the images in order to train and validate AI algorithms ready to recognize and classify hematopoietic stem cells. Chandradevan *et al*.^10^ and Wang *et al*.^11^ have independently developed an AI algorithm based on whole slide images, reaching recall levels higher than 90%. Matek *et al*. made a commendable effort by digitizing with a digital camera samples from 945 patients to develop an AI algorithm that identifies 21 cell classes^12^. Su *et al*.^13^ and Wu *et al*.^14^, used as well in their respective investigations a digital camera to digitize BMA samples. Tayebi *et al*., who also analyzed whole slide images, introduced an interesting concept, the so-called Histogram of Cell Types (HCT) quantifying bone marrow cell class probability distribution for each BMA sample^15^. Nevertheless, none of the above have considered how these tools are going to be implemented in an actual clinical practice that allows the interaction between the professional and the suggested results.

On the other hand, other authors have proposed a specially designed system for digitizing BMA samples with an automatic acquisition, scanning and optical focusing of the samples, and for automatic analysis of cell differential counts supported by machine learning models^16^. This system has been preliminary validated on 124 samples, and good agreement was reported between AI-based automatic cell counting and conventional manual counting of cell series proportions^17^.

Despite the effort to develop AI algorithms to assist BMA analysis, these previously proposed tools have not been widely implemented in real clinical environments due to the complexity, clinical workflow disruption, and the high price of the devices that allow the digitization of BMA samples, leaving BMA analysis as a manual, in-person, and synchronous process that has not been yet hit by the wave of digital transformation that has impacted other diagnostic tests.

Interestingly enough, the above-mentioned problems are to some extent shared with the analysis of peripheral blood (PB) samples, but they have been extensively addressed on numerous occasions and in commercial products^18,19^. The main difference that justifies this discrepancy in available solutions is due to the difficulty of obtaining high-quality, high-resolution images of a BMA sample which is thicker than a PB sample, with a more complex composition (mostly because of the presence of spikes, particles, and fat). In addition, as it requires scanning at 1000x magnification (or at least 400x) with immersion oil, it makes it more difficult to use commercial scanners that are usually employed for histology samples but are not valid for cytology and thicker samples like BMA smears.

With these previous considerations in mind, as a novelty compared to previous research in this field, we aimed to develop and evaluate an integrated digital system to cover the entire process, from BMA sample digitization to DCC facilitated by human-AI interaction, using a conventional optical microscope, a 3D printed device, a smartphone, a mobile application, and a web-based telemedicine platform which integrates the analysis from AI algorithms. By taking advantage of this holistic digital system, it could be possible to implement the solution in any hematology department in the world without incorporating specific and complex medical electronic devices into the clinical workflow.

## 2. Materials and Methods

### 2.1. System

The workflow of the AI-assisted digital DCC system is presented in **Figure 1**. The pipeline is as follows: first, the user digitizes a BMA sample using a 3D-printed microscopy arm that allows attaching a smartphone with a conventional optical microscope. The user uses a specific mobile app and digitizes at least 20 different fields of the sample using a 100x objective. The acquired images are automatically uploaded through the mobile app to a web telemedicine platform, where are subsequently processed by an AI algorithm that automatically detects and differentiates all nucleated cells in the sample into six possible cell lineages. The user can review AI predictions and confirm or edit them in case they disagree with the AI model. Lastly, cell series proportions are calculated generating the DCC.

**Figure 1.**
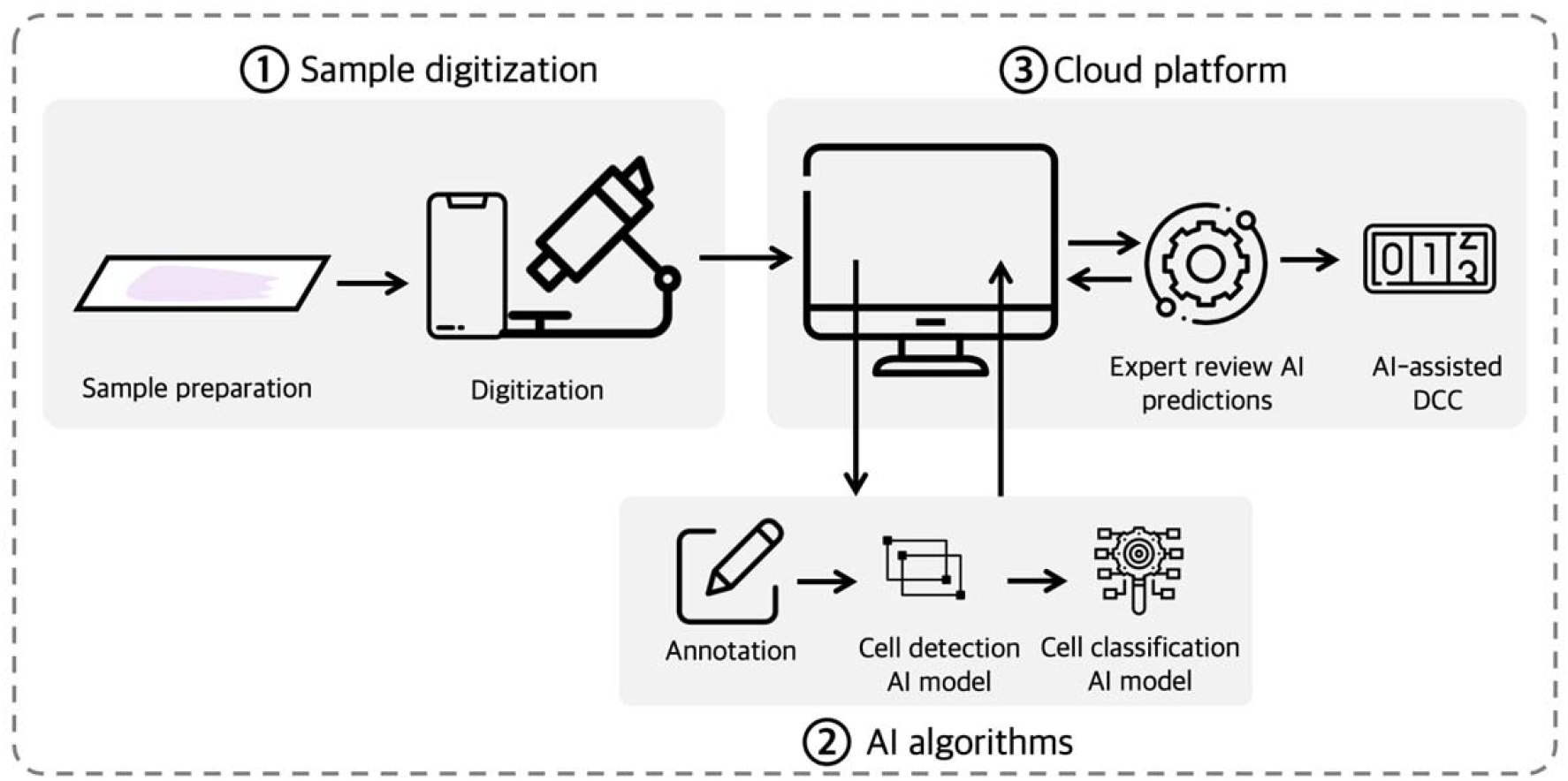
Workflow of the system, composed of three main components. (1) digitization, (2) AI algorithms and (3) cloud visualization platform.

#### 2.1.1. Digitization

To digitize BMA samples, the system includes a 3D-printed device that allows coupling and aligning a smartphone’s camera with a conventional optical microscope’s eyepiece lens^20^. In this way, this device converts any optical microscope into a digital one and can be used to obtain digital images of BMA samples. The smartphone uses a mobile app developed to acquire patient metadata and which was previously customized specifically for fast, standardized, and easy digitization of BMA microscopy images (see **Figure 2**).

**Figure 2.**
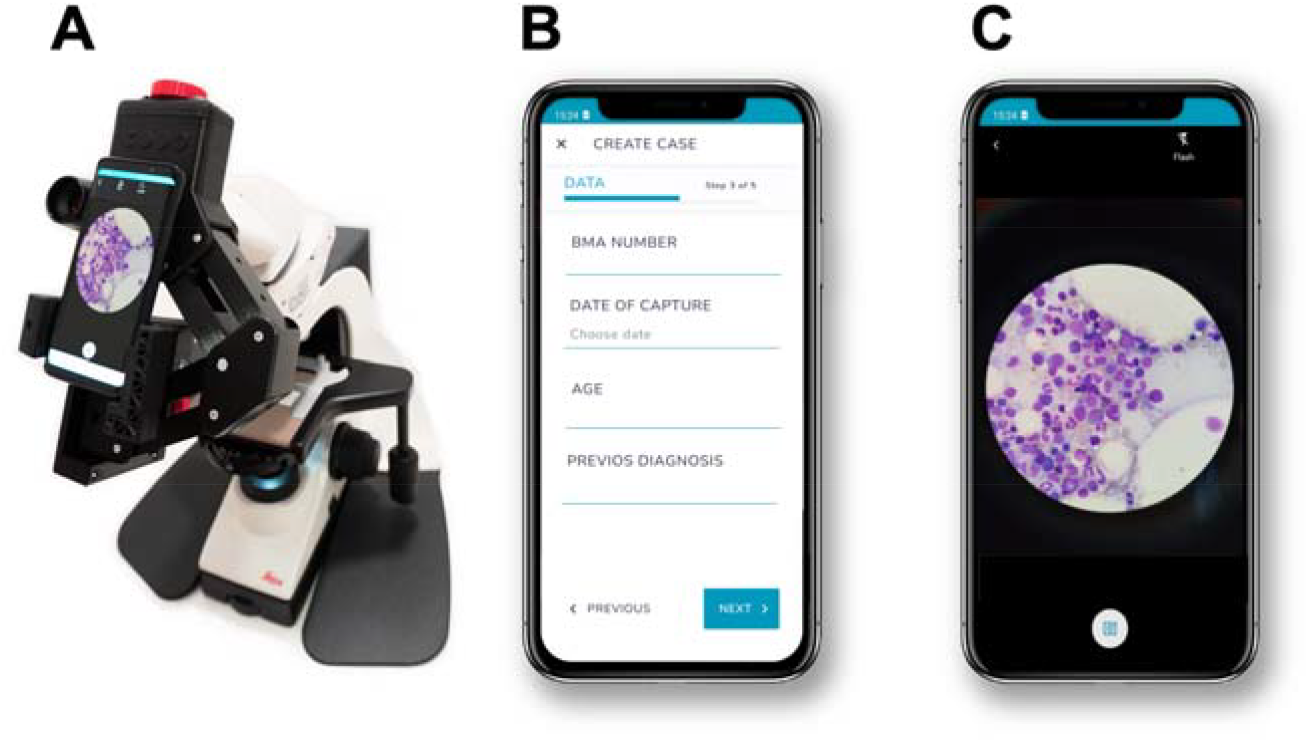
Sample digitization system. **A**: 3D-printed microscopy adapter arm that allows coupling a smartphone with any conventional microscope. **B** and **C**: Screenshots of the mobile app for standardized BMA image acquisition. The app allows collecting metadata associated to each BMA sample.

Users employed this digitization system to acquire photographs of 20 microscope fields for each BMA sample, ensuring that more than 500 cells were digitized. All acquired images were uploaded from the mobile app to a web-based telemedicine platform that allowed a remote visualization and analysis of BMA images. Patient data such as age and previous diagnosis were also collected through the mobile app and transferred to the telemedicine platform.

BMA samples used for training the AI algorithm were digitized using a BQ Aquaris X2 smartphone model, while samples used for the evaluation of the entire AI-assisted digital DCC system were digitized using two different smartphone models (BQ Aquaris X2 and Xiaomi Redmi Note 8T), to increase the variability in terms of mobile models, and reduce the possible bias of this variable. All images were acquired at an image resolution of 12Mpx. The smartphones were attached with the 3D-printed device to the ocular of a light microscope (Leica DM 2000 LED) and using a 100x objective (1000x total magnification).

#### 2.1.2. AI algorithm

The AI algorithm was developed for the automatic identification and differentiation of nucleated cells in BMA images into seven different classes: myeloid, erythroid, monocytic, lymphoid, blasts, plasma cells, and artifacts (cells without optimal cytologic characteristics). The algorithm consisted of a two-stage cell detection and lineage classification deep learning-based model. First, it detects all nucleated cells without regard to the cell lineage. The detected cells are subsequently introduced into the classification model, which can distinguish between different cell lineage classes.

Once an entire image representing a microscope field of view is processed by the AI algorithm, cell series proportions are calculated automatically for all the six lineage classes under study. The proportion of “artifact” class was not reported, as it has no clinical relevance.

#### 2.1.3. AI-assisted web telemedicine platform

All acquired images with the mobile app are transferred to the cloud telemedicine platform, where images can be visualized in an easy-to-use dashboard, allowing scrolling, and zooming the images. This platform also allows image labeling, which can be used by experts to digitally analyze BMA samples. In addition, these manually generated labels can be used for training the algorithms.

On the other hand, the developed AI algorithm is embedded into the platform so that all uploaded images are processed by the model, and AI-annotations are visualized. The platform includes a review terminal, which is designed for experts to review results produced by the AI and confirm or edit AI suggestions in case they disagree with them by either re-classifying mis-differentiated cells in the image or by labelling new undetected cells. Comparison between AI predictions and reviewed classification can be performed to assess AI model performance and usability of the system. This process is designed to speed up the diagnosis process as the user only must review and modify those cells with which they disagree, rather than manually differentiating and counting the required 500 cells per sample.

### 2.2. Study design and sample collection

Two different datasets were collected for this study. The first one was collected for the development of an AI algorithm for automatic identification and classification of hematopoietic stem cells in BMA samples. This dataset consisted of 101 BMA randomly selected samples that were retrospectively extracted between 2019 and 2021 at the Hematology Service of the University Hospital 12 de Octubre (Madrid, Spain). The age of included patients ranged from 1 to 87 (mean 57.5 years old).

Secondly, we validated the entire AI-based digital system for assisting hematologists perform a DCC. For this purpose, a second dataset was collected which was composed of 16 BMA samples that were extracted between December 2021 and February 2022 at the same center, with an age range from 2 to 82 years old (mean of 55.5 years). The BMA samples were selected considering that different hematological diseases should be represented to validate the algorithm. Patient characteristics of both study cohorts are shown in **Table 1**.

**Table 1.** Patient characteristics of the study cohort.

|  |  | Development of the<br>AI algorithm | System evaluation |
| --- | --- | --- | --- |
| Age | (mean ± std) (y) | 57.56 ± 15.71 | 55.5 ± 19.89 |
| Gender | Female | 57 (60.64%) | 9 (56.25%) |
|  | Male | 37 (39.36%) | 7 (43.75%) |
| Diagnosis | Acute lymphoblastic leukemia | 6 (6.38%) | 2 (12.5%) |
|  | Acute myeloid leukemia | 20 (21.28%) | 4 (25%) |
|  | Chronic lymphocytic leukemia | 2 (2.13%) | 1 (6.25%) |
|  | Chronic myeloid leukemia | 5 (5.32%) | 1 (6.25%) |
|  | Diffuse large B cell lymphoma | 4 (4.26%) | 1 (6.25%) |
|  | Follicular lymphoma | 5 (5.32%) | - |
|  | Multiple myeloma and other plasma cells dyscrasias | 33 (35.11%) | 5 (31.25%) |
|  | Other lymphomas | 7 (7.45%) | 1 (6.25%) |
|  | Reactive bone marrow | 5 (5.32%) | - |
|  | Others | 7 (7.45%) | 1 (6.25%) |

All BMA samples were prepared according to standard protocols and using May-Grünwald-Giemsa staining. We assessed sample preparation quality to discard those BMA samples without proper quality staining, insufficient lump, or those with a certain level of dysplasia.

Ethical approval for the study was obtained from the Hospital 12 de Octubre Ethics Committee for Research with medicinal products (Num. Ref. 20/430). This study was conducted in accordance with the Declaration of Helsinki, all BMA samples used in this study were anonymized according to local guidelines and informed consent was obtained from all subjects.

### 2.3. Development and training of AI algorithms

The cell labeling process for training the AI algorithms was performed using the web-based telemedicine platform, where expert hematologists manually annotated individual cells using a point-based annotation tool by placing a label point corresponding to one of the 7 possible classes (6 different cell lineages and artifact class) in the center of each cell. These annotated images were used for training and validating the classification algorithm. Additionally, and for training the cell detection algorithm, cells were annotated without regard to the cell class by manually drawing a bounding box around all cells present in a given image (microscope field). **Figure 3** shows an example of an annotation procedure for both cases (bounding boxes for training cell detection algorithm and point-based labels for training cell classification algorithm), which were both performed in the web telemedicine platform.

**Figure 3.**
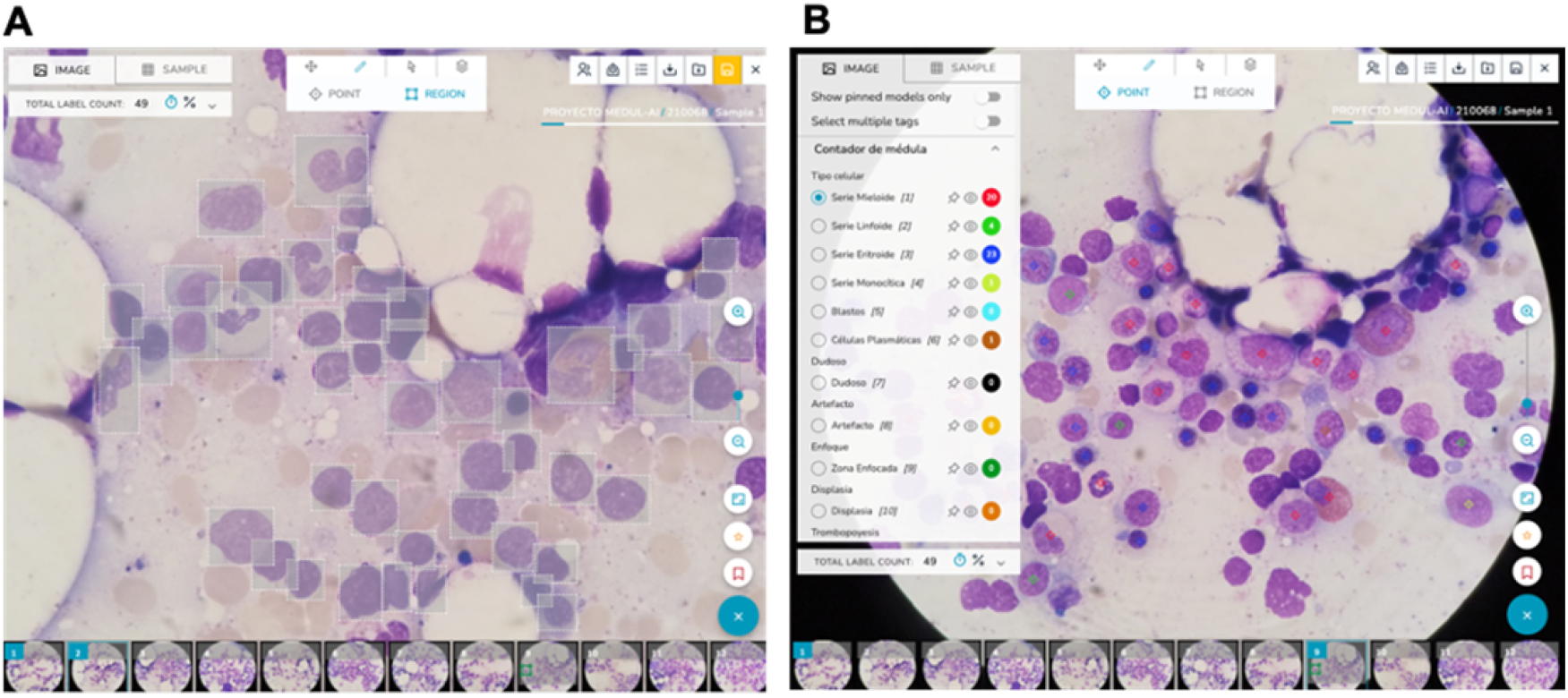
Screenshots of the web telemedicine platform for image labeling. **A**: Manually placed bounding boxes around all cells in a given image for training the detection algorithm. **B**: Point-based annotations for training the classification algorithm.

For training the classification algorithm, a total of 61,344 cells were identified and classified with one of the classes under study by a panel of 3 different hematologists (randomly selected from the 6 who participated in the study) according to 7 different classes: myeloid, erythroid, monocytic, lymphoid, blasts, plasma cells and artifact (cells without optimal cytologic characteristics). From these labels, and in order to train the algorithm only with those cells with full agreement among observers, we discarded 4,171 cells and thus, we only used 57,173 labels for training the algorithm, with the following distribution: myeloid (n=13,497), erythroid (n=7,164), monocytic (n=1,055), lymphoid (n=2,793), blasts (n=2,444), plasma cells (n=1,105) and artifact (n=29,115). From these 57,173 total labels, 80% was used for training the classification algorithm, while the remaining 20% was used for preliminary assessment of AI performance.

On the other hand, a total of 13,494 cells annotated in the form of bounding boxes were used for training (n=4,358) and validating (n=9,136) the detection algorithm.

The proposed algorithm for cell detection was based on the Single-Shot Detection (SSD)^21^ architecture together with the MobileNet V2^22^ backbone network. Both networks have been designed to be lightweight and computationally efficient, so they can be efficiently integrated into embedded vision systems, such as smartphones.

The detection network detected all cells appearing in a given image (microscope field) without regard to the cell lineage. The detected cells were subsequently processed by a classification algorithm for assigning them to one of the 7 possible classes.

Small image patches (200×200 pixels, 14.4×14.4 mm2) were extracted around the center of the bounding box of each detected cell, which were further classified by a Xception deep learning architecture^23^. Data augmentation including rotation, horizontal and vertical flips was included to make the classification algorithm robust and improve accuracy. Additionally, brightness, contrast and color modifications were included to mimic possible variations in variables such as microscope and smartphone used for sample digitization as well as possible variation in the sample staining procedure. No additional preprocessing step was performed in images to train the algorithms.

### 2.4. Evaluation of the system

For comparative purposes and to evaluate the entire AI-assisted digital DCC system, all 16 BMA samples were analyzed in the telemedicine platform by 4 different expert hematologists. For each sample preparation, one hematologist performed the digital analysis of the sample in a blinded fashion without the assistance of the AI (i.e., labeling and performing the DCC from scratch), while the other three hematologists analyzed the same sample assisted by the AI algorithm. All hematologists were asked to analyze and count at least 500 nucleated cells on each aspirate smear, following international recommendations on the diagnosis of hematology disorders^7,9^.

Hematologists who were assisted by the AI were presented with the predictions made by the algorithm and were asked to review each of the classified cells and re-label those incorrectly classified or undetected. Those hematologists who did not have the assistance of AI manually labeled from scratch at least 500 cells from each preparation.

The selection of the hematologists who were assisted by the AI and those who performed the blinded analysis was done on a rotating basis.

The relative percentages of each cell series for all preparations were calculated and compared for both methods (digital DCC and AI-assisted DCC).

The time needed to complete the analysis of a BMA sample when AI assistance was used, and when the analysis was performed in a blinded fashion was measured. Analysis times were compared to assess whether the AI-assisted system could reduce DCC diagnosis time. Additionally, as each of the BMA samples was analyzed by 4 experienced hematologists, we also quantified the inter-observer variability by measuring the agreement in the cell classification among different experts.

Lastly, we also assessed the performance of the AI algorithm on these 16 BMA independent samples by comparing the AI predictions against a consensus labeling defined as the majority among the 4 experts for each cell.

## 3. Results

From the 61,344 cells manually identified and classified along the 101 BMA samples collected for algorithm development, a total of 5,144 cells were separated for preliminary performance evaluation (20% of the available images). Cells annotated as artifact were not included in this analysis.

On the other hand, each of the 16 BMA samples for evaluating the entire system were analyzed by 4 different experienced hematologists. Each expert identified and classified at least 500 cells for each sample. Experts placed 7,840, 7,922, 7,908 and 8,375 labels respectively along the 16 BMA samples. From these labels, only 4,401 corresponded to the same cell, so that each of these cells was classified by the 4 experts, in addition to the AI algorithm.

### 3.1. Performance of the AI algorithm

The detection algorithm achieved a high overall accuracy for detecting BM nucleated cells in microscopy images, with sensitivity of 91.6% (95%CI 90.6%-92.7), and precision of 91.3% (95% CI 90.2%, 92.3%).

Two data sets were used to evaluate the performance of the classification model. First, the performance was evaluated on the validation set (20% of the training set) which included the above-mentioned 5,144 cells. The algorithm was also evaluated on an independent test set obtained from the 16 BMA samples used for evaluating the entire system comprising 4,401 cells classified by 4 different experts, where the ground-truth was established using the majority voting rule (consensus) among the 4 experts. The total validation set consisted of 9,545 cells. **Table 2** shows the average performance of the model in the total validation set as well as detailed performance for each cell lineage class for both validation sets independently. The confusion matrix for the total validation set (9,545 cells) is shown in **Figure 4**. The overall accuracy was 92.97%, although it is worth noting the decrease in performance in those classes with fewer training images, such as monocytic and plasma cells. However, this fact is easily remedied by increasing the number of training images for these classes.

**Table 2.** Performance of the AI algorithm for detecting and classifying cells in BMA samples. N: number of cells used for evaluating the performance.

|  | 20% validation set |  | Independent validation set |  | Total validation |  |  |
| --- | --- | --- | --- | --- | --- | --- | --- |
|  | Accuracy | N | Accuracy | N | Accuracy | 95% CI | N |
| Lymphoid | 86.56 | 424 | 86.82 | 258 | 86.66 | [ 84.10 - 89.21 ] | 682 |
| Erythroid | 96.18 | 1284 | 92.43 | 1030 | 94.51 | [ 93.58 - 95.44 ] | 2314 |
| Myeloid | 98.17 | 2565 | 98.25 | 2286 | 98.21 | [ 97.83 - 98.58 ] | 4851 |
| Monocytic | 56.10 | 205 | 67.63 | 139 | 60.76 | [ 55.60 - 65.92 ] | 344 |
| Blasts | 87.42 | 469 | 82.60 | 638 | 84.64 | [ 82.52 - 86.77 ] | 1107 |
| Plasma cells | 79.70 | 197 | 58.00 | 50 | 75.30 | [ 69.93 - 80.68 ] | 247 |
| **Average** | **93.35** | **5144** | **92.52** | **4401** | **92.97** | **[ 92.46 - 93.48 ]** | **9545** |

**Figure 4.**
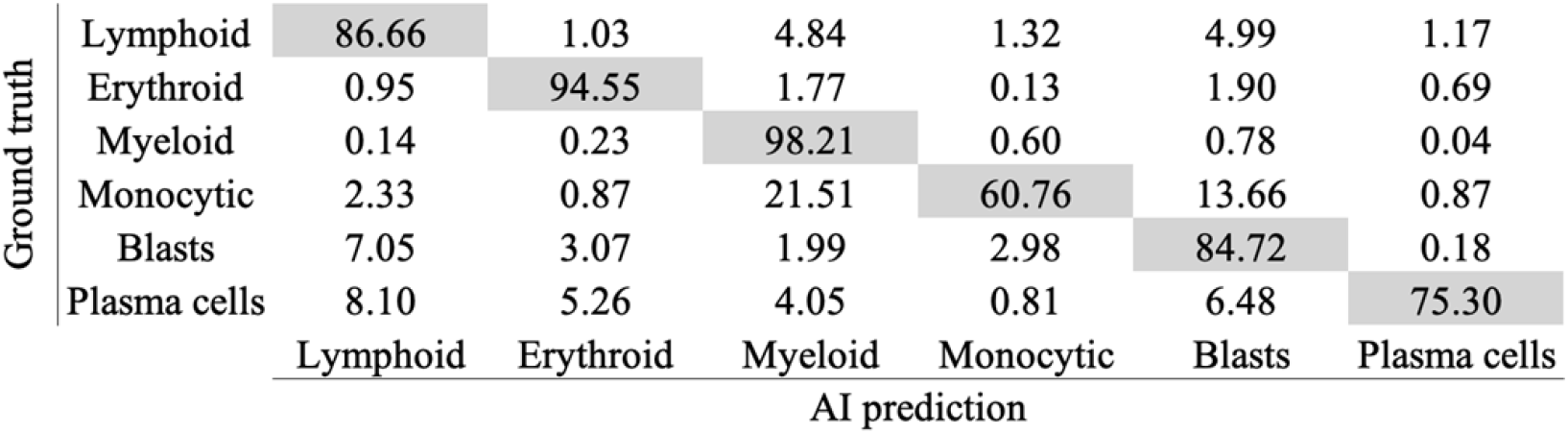
Performance evaluation of the AI algorithm. Confusion matrix comparing ground truth and AI predictions.

### 3.2. Evaluation of the system

#### 3.2.1. Interobserver variability

BMA analysis presents a high level of interobserver variability when differentiating hematopoietic stem cell lineages. Cohen’s kappa score was used to evaluate interobserver agreement in the classification of cell lineages. The agreement was performed on the 4,401 BM cells annotated by 4 experts. **Table 3** presents the kappa results for all pairs of the observers as well as for the agreement between each observer and the AI predictions. Mean interobserver agreement was 0.895. Similar agreement was found when comparing experts and AI predictions, with a mean kappa score of 0.871.

**Table 3.**
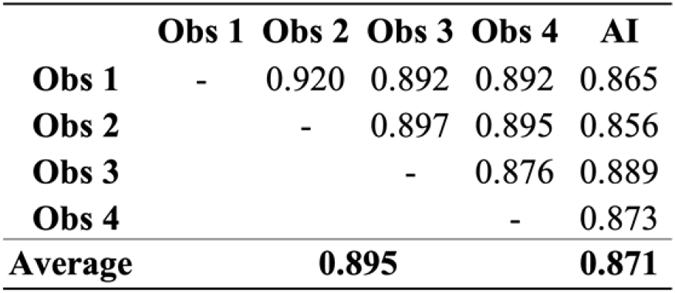
Cohen’s kappa score representing interobserver agreement and agreement between observers and AI predictions.

On the other hand, and to assess whether the assistance of the AI can reduce the interobserver variability among hematologists when analyzing BMA samples, we have computed interobserver agreement on those samples that were analyzed in a blinded fashion, as well as on samples where hematologists were assisted by the AI. **Table 4** shows the comparison between both interobserver agreement values (blinded and AI-assisted analysis) and, as it can be seen, interobserver agreement is considerably increased when AI is used, from 0.88 to 0.93. The difference in interobserver agreement between both methods is statistically significant (p-value<0.0001).

**Table 4.**
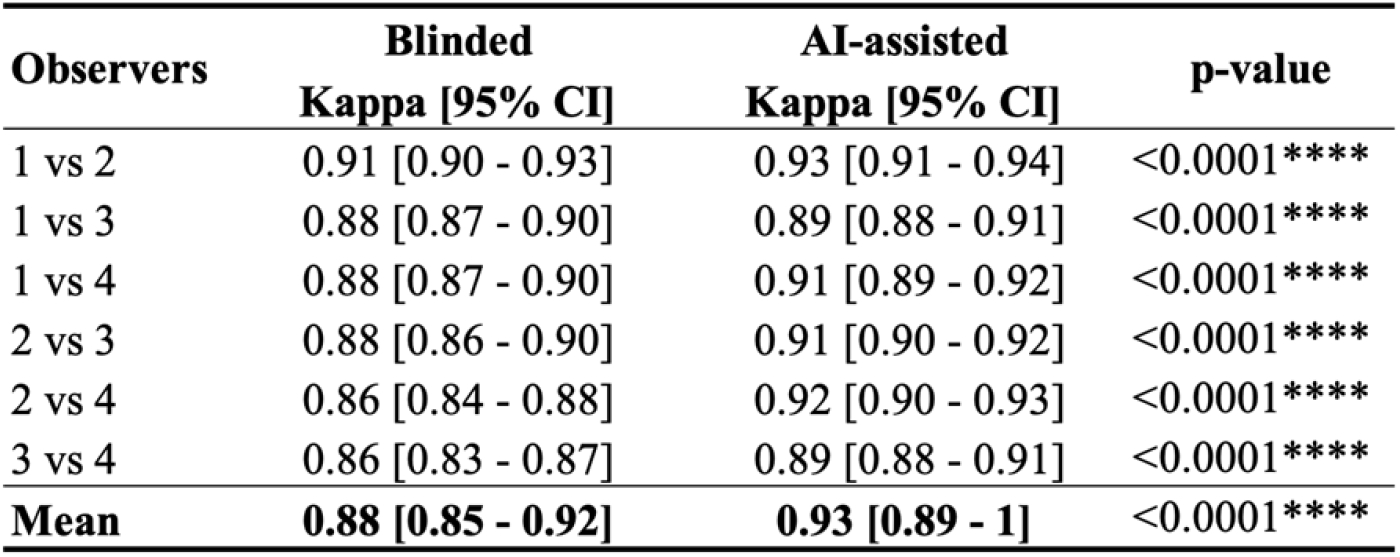
Interobserver agreement when BMA samples are analyzed in a blinded fashion (left) and when hematologists are assisted by AI (right).

#### 3.2.2. Analysis time

To assess whether the AI assistance can reduce analysis time, we have measured the time needed to complete a BMA analysis when users were assisted by AI and when they had to perform the analysis from scratch (i.e., without the assistance of AI).

**Table 5** summarizes the average time needed to analyze a single cell for each of the 4 involved hematologists, as well as the time needed to complete an entire BMA analysis, which usually comprises the identification of at least 500 cells. As derived from the table, it is shown that when users used the AI assistance, the analysis time was reduced by a factor of 18.75%.

**Table 5.**
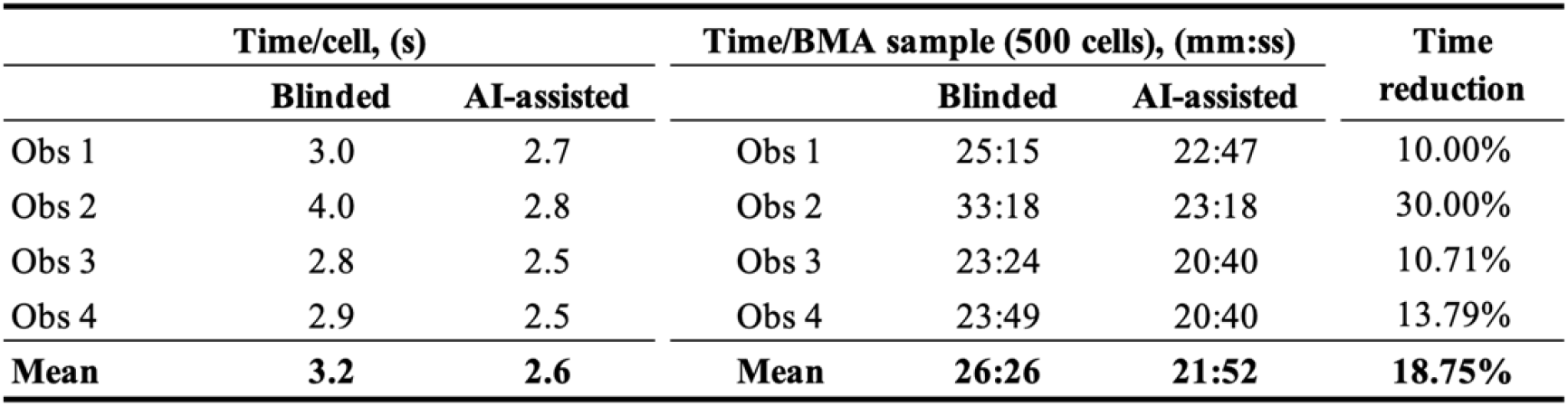
Comparison of the time needed to complete an analysis of a single cell (left) and a whole BMA sample (right) when the user used the AI assistance and when the analysis was performed in a blinded fashion. Note: mm: minutes; s/ss: seconds.

#### 3.2.3. Usability assessment

As part of the objectives of this work, we wanted to evaluate the usability of the proposed AI-assisted digital DCC system. For this purpose, we designed an anonymized usability survey that was completed by the four hematologists who used the proposed system. The participants showed a high level of satisfaction in using the system (average score of 4.59 out of 5) and considered it a useful tool in their daily work. Additionally, the results derived from the usability survey show that the automatic hematopoietic stem cell lineage percentages predicted by the AI algorithm were sufficiently reliable, even though some cells were misclassified and had to be modified in the review process.

## 4. Conclusions and discussion

We have presented a digital system that allows counting cell types from BMAs. The proposed system not only provides an accessible and simple way to digitize, store and label BMAs remotely but also is supported by an AI pipeline which speeds up the time required for the analysis and reduces the interobserver variability. It proposes therefore a very concrete analysis workflow which integrates the AI algorithm with the clinical practice in a regular hospital workflow.

Digitizing BMAs has some direct implications as it is possible to easily examine the samples remotely, to share it for second opinions or to analyze samples from the same patient over time. Furthermore, it allows precise quantitative analysis and research of cell types and morphologies, as well as to test new clinical hypotheses. Some clinical interpretations (clinical remissions and response to treatment) rely on an expert analysis of a limited number of fields or cells and with the proposed system working in an optimized manner it will be possible to analyze a higher number of cells without increasing the time of analysis, hence being able to measure with more precision.

The study presented in this work has required the creation of an extensive database for training an algorithm able to classify different cell types. It is worth mentioning that all the samples used come from one single hospital, and though the bias depending on sample preparation is limited, a further multi-centric validation is suggested to understand the bias introduced by variability of the preparation. It is also worth pointing out that the digitization process enabled by mobile phones has standardized acquisition parameters available in any android smartphone and results in no difference between smartphones.

Digitalization based on a smartphone without the requirement of scanners and complex equipment lowers barriers to scale all the benefits of digitalization of BMAs including accuracy, possibility to share and do remote analysis and reproducibility.

Once data is digitized and considering real life systems that go beyond AI algorithms that work in isolation, we show a system with all the required components: in this case not only a two-stage algorithm, but a visualization and interaction tool to integrate human and AI support. The current challenge of AI in the medical field has to do with the integration of AI systems in the clinical workflows and for this purpose technologists should design systems that optimize clinical outcomes, being multidisciplinary projects by nature.

Future work includes improving accuracy of the AI pipeline, including new cell types, adding other biomarkers and the unsupervised discovery of relevant patterns and cell morphologies correlated with other types of analysis including genetic data. From a technical perspective, we could foresee an algorithm with higher levels of precision—that is just a question of enough training data— allowing almost fully automated counting. Further research in ways to integrate such knowledge and human-AI feedback loops will be required. From cell counting to disease prediction, digitalization can be key to unlock a big potential for hematology.

## Data Availability

For data sharing requests, please contact Miguel Luengo-Oroz

## Acknowledgments

This project has received funding from the European Union’s Horizon 2020 research and innovation program under grant agreement No 881062. DB-P was supported by grant PTQ2020-011340/AEI/10.13039/501100011033 funded by the Spanish State Investigation Agency. RG-V holds a Formación de Profesorado Universitario (FPU19/04933) grant from the Ministry of Science, Innovation and Universities of Spain Government. AR-G holds the FEHH 2021 research grant from the Spanish Society of Hematology. LL was supported by a predoctoral grant IND2019/TIC-17167 (Comunidad de Madrid, Spain).

## Authorship

DB-P, SRC, AMU, EA, ED, MP, AS, MJL-C, RAD, JM-L, ML and MLO conceived and designed the study. MGR, RT-M, AB-F, MHS, RG-V, MLM, AR-G, AO-R, ABS contributed, digitized, and analyzed the samples. DB-P, LL, DC, AV, JG-V designed and developed the software. DB-P, SRC and MLO analyzed and interpreted the results. DB-P and SRC wrote the main manuscript text. All authors interpreted the results and revised and edited the manuscript.

DB-P, SRC, AMU, EA, LL, ED, DC, MP, AV, JG-V, MJL-C, AS, ML and MLO hold shares or phantom shares of Spotlab. The rest of the authors declare no conflict of interest.

## Notes

### Funding Statement

This project has received funding from the European Union Horizon 2020 research and innovation program under grant agreement No 881062. DB-P was supported by grant PTQ2020-011340/AEI/10.13039/501100011033 funded by the Spanish State Investigation Agency. RG-V holds a Formacion de Profesorado Universitario (FPU19/04933) grant from the Ministry of Science, Innovation and Universities of Spain Government. AR-G holds the FEHH 2021 research grant from the Spanish Society of Hematology. LL was supported by a predoctoral grant IND2019/TIC-17167 (Comunidad de Madrid, Spain).

### Author Declarations

Ethical approval for the study was obtained from the Hospital 12 de Octubre Ethics Committee for Research with medicinal products (Num. Ref. 20/430). This study was conducted in accordance with the Declaration of Helsinki, all samples used in this study were anonymized according to local guidelines and informed consent was obtained from all subjects.

