## Supplementary Material for "Digital system augmented by artificial intelligence to interpret bone marrow samples for hematological disease diagnosis"

Supplementary material: Table 1S

| **Reference** | **Digitization** | **Dataset** | **Classes** | **Annotators** | **AI performance** |
|---|---|---|---|---|---|
| (Chandradevan et al., 2020) | Aperio T2 40x | -17 patients - >10000 nonneoplastic cells<br>- 2 MM and 3 AML (validation) | 13 | 3 pathologists | - Detection (0.959 ± 0.008 precision-recall AUC)<br>- Classification (0.982 ± 0.03 ROC AUC) |
| (Choi et al., 2017) | 1000x light microscope, camera? | - 30 BMA samples from 10 patients (2174 cells) | 10 (including only erythroid and myeloid subtypes) | 2 hematologists | - Accuracy 97.06%<br>- Precision 97.13%<br>- Recall 97.06%, - F-1 score 97.1% |
| (Fu et al., 2020) | Morphogo WSI 400x | - 230 BMA samples | 12 | 2 pathologists consensus | - Accuracy above 85.7% for all cell lineages<br>- Sensitivity 69.4%<br>- Specificity 97.2%<br>- Correlation (AI Vs. 2 pathologists consensus) r ≥ 0.762 |
| (Jin et al., 2020) | Morphogo WSI | - Development: 3000 BMA (>600,000 cells)<br>- Validation: 124 BMA (>31,000) | 11 | Number of pathologists not specified | - Accuracy 90.1%<br>- Intraclass correlation coefficient (ICC): Average ICC ≥ 0.883; Granulocytes 0.893; |

| | | | | | |
|---|---|---|---|---|---|
| | | | | | Erythrocytes 0.883;<br><br>Lymphocytes 0.763;<br><br>Monocytes 0.449;<br><br>Plasma cells 0.368. |
| (Matek et al., 2021) | CCD camera mounted on a brightfield microscope (Zeiss Axio Imager Z2) | - 945 patients, (171 374 cells) | 21 | Morphologists (number not specified) | - Precision and recall, respectively: Band neutrophils 0.54 and 0.65; Segmented neutrophils 0.92 and 0.71; Lymphocytes 0.90 and 0.70; Monocytes 0.57 and 0.70; Eosinophils 0.85 and 0.91; Basophils 0.14 and 0.64; Metamyelocytes 0.30 and 0.64; Myelocytes 0.52 and 0.59; Promyelocytes 0.76 and 0.72; Blasts 0.75 and 0.65; Plasma cells 0.81 and 0.84; Proerythroblasts 0.57 and 0.63; Erythroblasts 0.88 and 0.82; Hairy cells 0.35 and 0.80; Abnormal eosinophils 0.02 and 0.20; Immature lymphocytes 0.08 and 0.53; Faggot cells 0.17 and 0.63. |
| (Su et al., 2021) | Digital microscopy camera | - 230 BMA images, 8239 cells | 8 | - Expert hematopathologists (number not specified) | - Cell detector precision: >83%<br><br>- Average cell classification precision: 0.9530, and recall 0.9576 |

| | | | | | |
|---|---|---|---|---|---|
| (C.-W. Wang et al., 2022) | WSI at 400x magnification (device specification not available) | - 12426 cells<br>- External validation: 3005 cells | 16 | Not available | - Recall and accuracy of 0.905 ± 0.078 and 0.989 ± 0.006, respectively<br>- External validation: Recall and accuracy of 0.842 and 0.988, respectively |
| (D. Wang et al., 2021) | Not specified | - 609 cells | 7 | Not specified | Up to 98.8% accuracy (not clearly specified) |
| (Wu et al., 2020) | 1000x microscope and digital camera | -Development: 42 samples (>17,000 cells)<br>- Validation: 70 samples Leukemia and MDS)<br>- Competition: 10 samples | 8 | 3 hematologists | - Cell detection average prediction 67.4%<br>- Validation: leukemia >5% blasts AUC 0.948, Leukemia >20% blasts AUC 0.942, MDS >5% blasts AUC 0.888, MDS>20% blasts AUC 0.762<br>- Competition: correlations between BMSNet and flow cytometry 0.960 |
| (Tayebi et al., 2022) | WSI acquired with Aperio Scanscope AT<br>Turbo and | - 250 patient WSI for the development of the ROI detection model | 19 | - Expert hematopathologist s (number not specified) | - Region detection average accuracy of 97%.<br>- Cell detection and classification average precision of 75%. |

| | | |
|---|---|---|
| | Huron TissueScope at 40X | - 500 individual patients' WSI (26,782 cells) |

**Table 1S**. Literature review of AI algorithms previously proposed for the analysis of BMA samples.
